# Efficacy and safety of levetiracetam versus phenytoin as second line antiepileptic agent in Pediatric convulsive status epilepticus: A systematic review and meta-analysis of randomized controlled trials

**DOI:** 10.1101/2020.10.29.20222133

**Authors:** Suresh Kumar Angurana, Renu Suthar

## Abstract

**Objective:** To evaluate the efficacy and safety of Levetiracetam (LEV) in comparison to phenytoin (PHT) as second line antiseizure medication (ASM) for Pediatric convulsive status epilepticus (SE).

**Data source:** PubMed, Embase, Google scholar, Scopus, Cumulative Index to Nursing and Allied Health Literature (CINAHL), Cochrane Database of Systematic Reviews and Cochrane Central Register of Controlled Trials.

**Study selection:** Randomized controlled trials (RCTs) assessing LEV and PHT as second line agent for convulsive SE in children <18 years published between 1^st^ January 2000 to 30^th^ September 2020.

**Data extraction:** The data was pooled regarding the proportion of children achieving seizure cessation within 5-60 minutes of completion of study drug infusion (primary outcome); and seizure cessation within 5 minutes, time to achieve seizure cessation, seizure recurrence between 1-24 hours, intubation, and cardiovascular instability (secondary outcomes). Data was analysed using RevMan version 5.4 and quality analysis was done using Cochrane risk-of-bias tool. The study protocol was submitted to PROSPERO for registration.

**Data synthesis:** Eleven RCTs with 2177 children (1024 received LEV and 988 received PHT) were enrolled. Seizure cessation within 5-60 minutes was similar with both the drugs [81% in LEV vs. 76% in PHT, risk ratio (RR)=1.04, 95% CI 0.9-1.13, p=0.29]. Seizure recurrences within 1-24 hours was higher with PHT as comparison to LEV (15% vs 9%, RR=0.64, 95% CI 0.42-0.99, p=0.04). Seizure cessation within 5 minutes, time to achieve seizure cessation, requirement of intubation/mechanical ventilation, and cardiovascular instability were similar with both the drugs. Three studies had low risk of bias and eight studies had high risk of bias.

**Conclusion:** The efficacy and safety of LEV is not superior to PHT as second line ASM medication for Pediatric convulsive SE. However, the seizure recurrences between 1-24 hours were significantly higher with PHT in comparison to LEV.

## Introduction

Status epilepticus (SE) is the most common neurological emergency in children. Convulsive SE is defined as continuous seizure activity of >30 minutes or ≥2 sequential seizures without full recovery of consciousness in between (1). For management purpose, seizures lasting for >5 minutes or child presenting seizing in emergency are managed as SE to reduce the risk of progression, neuronal damage, and consequences associated with prolonged seizure (1, 2). The overall incidence of Pediatric SE is about 20/100,000 children/year, with highest incidence among children <2 year of age, and common etiologies include febrile status and remote symptomatic seizures (1, 3-6). Convulsive SE is associated with mortality of 3-9% (3, 7, 8). The risk factors mortality in Pediatric SE include acute symptomatic etiology, age <5 years, prolonged and refractory SE, and generalized or multifocal electroencephalographic discharges (9, 10).

The goal of therapy for SE include initial stabilization of airway, breathing and circulation followed by seizure control, and identifying and treating the underlying etiology. Several guidelines have been published for uniform and time-bound management of Pediatric SE recommending intravenous benzodiazepines (diazepam, midazolam and lorazepam) as first line antiseizure medications (ASM) with maximum of two doses within 5-15 minutes followed by (fos)phenytoin/phenytoin (PHT), valproic acid, and levetiracetam (LEV) with in next 20-40 minutes as the second line ASM (2, 11, 12). The third line ASM include intravenous midazolam infusion, thiopentone coma, or anaesthetic agents in next 40-60 minutes (2, 11, 12). Patients requiring third line ASM often require mechanical ventilation and PICU admission. However, there is lack of consensus on use of second and third line ASM, and preferred anaesthetic agent (13).

Over the years, intravenous PHT remained the preferred second line ASM for Pediatric SE. However, PHT is associated with various adverse events including cardiac arrhythmia, hypotension, Steven Johnson syndrome, pancytopenia, and purple glove syndrome due to extravasation injury. However, Fosphenytoin, a prodrug of PHT, has advantages of lack of local reaction, phlebitis, and cardiotoxicity; and it can be infused rapidly (14). LEV is emerging as an alternative second line ASM for management of Pediatric SE with efficacy similar to PHT; and has better safety profile, rapid onset of action, ease in administration, require less intense monitoring; and better drug interaction, and pharmacokinetic profile (15-23).

The previous systematic reviews including adult and Pediatric SE cases reported efficacy of LEV similar to PHT, valproate, and lorazepam (24-28). However, recently few RCTs compared the safety and efficacy of LEV and PHT in Pediatric SE (15, 17-19). Therefore, in view of recent development of this evidence, we planned this systematic review and meta-analysis to study the efficacy and safety of LEV in comparison to PHT as second line ASM in management for Pediatric SE.

## Methodology

All RCTs involving children and adolescents from 1 months to 18 years of age with benzodiazepine refractory SE; treated with LEV and phenytoin/ fosphenytoin were included in this systematic review and meta-analysis as per the following criteria:

### Inclusion criteria

1. Age: 1 month to 18 years of age.
2. Diagnosis of convulsive SE refractory to benzodiazepines/established SE.
3. RCTs comparing standard therapy (phenytoin or fosphenytoin) with LEV.
4. Reported outcomes showing efficacy (seizure control) and safety profile (requirement of intubation, mechanical ventilation, and vasoactive drugs).

### Exclusion criteria

1. RCTs involving neonates <1 month of age and adults >18 years.
2. Observational studies.
3. Case reports.
4. Editorials, commentaries, reviews, viewpoints.
5. Dissertations and conference reports.
6. Narrative or systematic review.
7. RCTs published in languages other than English.

This systematic review was conducted as per the meta-analysis of randomized controlled trials guidelines (29). The protocol was submitted for registration in Prospero (on 1^st^ October 2020).

#### Search strategy

Two investigators (SAK and RS) performed independent literature search in electronic databases including PubMed, Embase, Google scholar, Scopus, Cumulative Index to Nursing and Allied Health Literature (CINAHL), Cochrane Database of Systematic Reviews and Cochrane Central Register of Controlled Trials data bases for studies published in English language from 1^st^ January 2000 to 30^th^ September 2020. The combination of the following keywords was used as the search strategy for literature search:

- Status epilepticus (SE), Convulsive SE, Pediatric SE, Established SE.
- Children aged >1 month and <18 years.
- Levetiracetam, phenytoin, fosphenytoin.
- Seizure control, seizure recurrence, mechanical ventilation, inotropes/vasoactive drugs.
- Articles, trials, randomized controlled trials, phase 3, phase 4 trials.

The references of included studies, review articles, and systematic reviews were screened. Articles published in the English language were included. The Preferred Reporting Items for Systematic Reviews and Meta-analyses guidelines were followed (23).

#### Study Selection

Both the investigators (SAK and RS) independently reviewed the titles and abstract for the eligibility criteria. Both the investigators (SKA and RS) examined the full articles and supplementary content for the exclusion and inclusion criteria. RCTs reporting comparison of LEV with phenytoin or fosphenytoin, seizure control, and adverse events were included. Studied involving both adult and pediatric cases was included if the information is separately available for the parameters of interest for children aged <18 years.

### Data extraction

A pre-designed standardized proforma was used for data extraction. Two investigators (SKA and RS) reviewed the full text of the papers/studies with the supplementary data (if available) from the eligible studies. Authors were requested to provide the data in the studies where the desired data was not separately presented in the paper. The data collection included first author’s name, journal name, year of publication, country, study design, number of centres, number of cases <18 years of age, benzodiazepines use, ASM used, dose of LEV and fos/phenytoin, time of study drug infusion, time to achieve seizure cessation, proportion of patients with seizure cessation, seizure recurrence, use of additional ASM, and adverse effects (requirement of vasoactive agents, intubation and mechanical ventilation).

Any disagreement at any point between the two investigators was sorted out through mutual discussion and consensus. To avoid duplicity of the data, efforts were be made to screen full text of all included studies for author names, setting, location, date and duration of the study, number of participants, and baseline data; and the studies with duplicate data will be excluded.

#### Quality assessment

The quality of included studies was assessed using the Cochrane risk of bias tool in the RevMan 5.4 software. The overall risk of bias to each included study was independently assessed by two investigators (SKA and RS). The studies were rated as either having low, unclear, or high-risk of bias.

#### Main outcomes

The main outcome was to study the efficacy and safety of LEV in comparison to PHT in children and adolescents <18 years with convulsive SE. The following discrete/continuous outcomes were collected and compared in 2 groups (LEV and PHT):

1. Number of children achieving cessation of seizures within 5 minutes of completing the study drug infusion.
2. Number of children achieving cessation of seizures within 5 minutes to 1 hour of completing the study drug infusion.
3. Number of children with seizure recurrence between 1-24 hours of completing study drug infusion.
4. The time (minutes) to achieve seizure cessation after completing study drug infusion (reported as mean and standard deviation).
5. Number of children requiring RSI/intubation/mechanical ventilation.
6. Number of children with cardiovascular instability (including requirement of vasoactive drugs, hypotension, and cardiac arrhythmias).

#### Data Synthesis

The initial data entry was done in a structured proforma and transferred to the Microsoft Excel 2020 (Microsoft, Redmond, WA). The descriptive analysis was performed using SPSS version 23 (IBM Corp. Released 2015. IBM SPSS Statistics for Windows, Version 23.0. Armonk, NY: IBM Corp). The data presented as number and percentages for categorical variables and mean or median (IQR) for continuous variables.

The meta-analysis was performed using Review Manager (RevMan, Computer program, version 5.4, The Cochrane collaboration, 2020) and results are presented as number, percentages, and risk ratio with 95% confidence interval for categorical outcome; and mean for continuous outcome. The data from the individual studies was pooled and analysed utilising random effect model with the assumption that the frequency of various parameters will be variable across the studies. The statistical heterogeneity among studies was be assessed by Chi-Square and I^2^ statistics. The studies were considered heterogeneous if I^2^ >50% and p<0.1.

## Results

A search according to the strategy revealed 450 studies, and 11 RCTs were included in the final analysis (Figure 1 and Table 1). Two RCTs were double-bind response adaptive trials (15, 16), one was double blind (19), and eight were open-label RCTs (17, 18, 20, 30-34). Nine RCTs included only Pediatric patients whereas two RCTs included both Pediatric and adult patients (15, 16), wherein data of Pediatric patients was reported separately in one RCT (15), and data was retrieved for one RCT (16). The studies were conducted in emergency department (ED) in nine RCTs and PICU in two RCTs (19, 33). Six RCTs were conducted in India (19, 20, 31-34), two in USA (15, 16), one each in Australia and New Zealand (17), UK (18), and Pakistan (30). Four RCTs were multicentric (15-18) and seven were conducted at single centre (19, 20, 30-34). The age range of the Pediatric only studies varied and included children and adolescents <18 years of age, and infants >1 month of age. The definition of SE also varied among the RCTs, eight RCTs explicitly mentioned benzodiazepine refractory SE or established SE (15-20, 30, 32) (Table 1).

**Figure 1:**
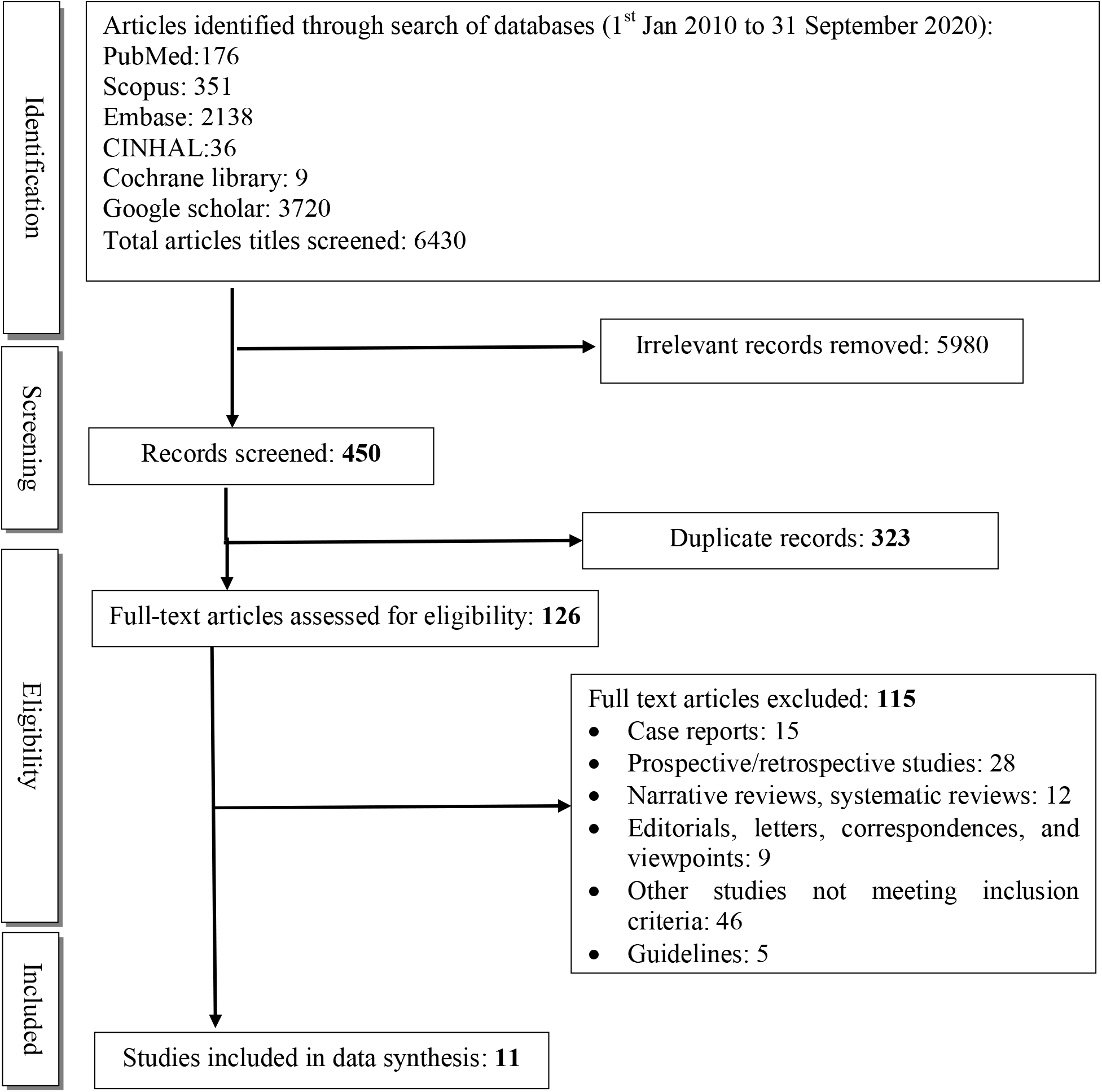
Flow diagram showing the process of study selection.

Among 11 included RCTs, a total of 2177 children <18 years were included, and 57% were male. Among the enrolled children, 31% (486/1559) were known epileptic and 34% (521/1509) had febrile SE. In 8 RCTs, only LEV and PHT were used, and in three RCTs, valproate was also used (15, 16, 19). A total of 1024 children received LEV and 988 children received PHT. The (Fos)phenytoin was uniformly used at 20 mg/kg dose in all the RCTs, while the dose of LEV was highly variable, ranging from 20 mg/kg to 60 mg/kg (15,16, 31-33). (Fos)phenytoin infusion was given over 10-20 minutes with strength of 1 mg/ml, while levetiracetam was infused intravenously at variable duration (range 5 min to 20 minutes). Mean duration of ongoing SE before initiation of the study medication was 30±23 minutes, (range 10-72 minutes).

### Efficacy analysis

The primary outcome used to assess the efficacy of LEV and PHT varied significantly among 11 RCTs. Three different types of outcome parameters have been used, time to achieve seizure cessation; proportion of subjects achieving seizure cessation after the completion of study drug infusion within certain time frame such as 5, 20, 30, 60 and at 120 minutes; and seizure recurrence upto 24 hours of study drug infusion and requirement of third line/additional AEDs. We analysed the efficacy outcome under four categories; studies reporting seizure cessation within 5 minutes of study drug infusion, studies reporting seizure cessation within 5 to 60 minutes of study drug infusion, studies reporting seizure recurrence between 1 hour to 24 hours of study drug infusion, and time to achieve seizure cessation.

#### Seizure cessation within 5 minutes (Figure 2)

Three RCTs (17, 32, 34) reported seizure cessation within 5 minutes of completion of study drug infusion. Meta-analysis of these three RCTs showed seizure cessation achieved in 59.6% (117/196) in LEV group and 65.4% (125/ 191) within 5 minutes of completion of study drug infusion. The treatment success rate was similar in both the groups with risk ratio of 0.96 (95% CI: 0.81-1.15, p=0.68). There was no statistical heterogeneity within the studies (I^2^ value of 41%, p=0.18) (Figure 2). The seizure cessation assessment was clinical, by most experienced physician at bedside which was defined as cessation of all forms of convulsive movements. Persistent seizures were defined as presence of either increased tone, jerking movements, nystagmoid eye movements, and reduced level of consciousness. The infusion of LEV was given over 5 minutes by Dalziel et al (17), 10 min by Wani et al (34), and 7 minutes by Senthilkumar et al (32).

**Figure 2:**
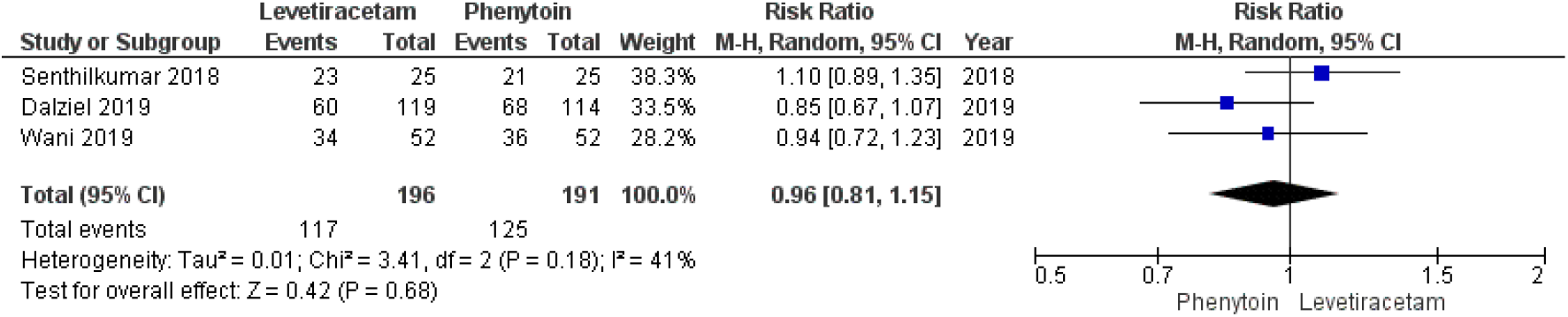
Seizure cessation with-in 5 minutes of completion of study drug infusion.

#### Seizure cessation within 60 minutes (Figure 3)

Eight RCTs reported achievement of seizure cessation within 60 minutes of completion of study drug infusion (15, 16, 18-20, 30, 34). In the meta-analysis, 81% (675/830) children in LEV group and 76% (617/808) in the PHT group achieved seizure cessation within 5 min to 60 minutes of completion of the study drug infusion [RR 1.04 (95% CI 0.9-1.13), p=0.29]. The RCTs included showed marked statistically heterogeneity (I^2^ value of 72% and p=0.0007) (Figure 3). Seizure cessation was assessed by Vignesh et al (19) at 15 minutes, Nalisetty et al (20) at 20 minutes, Sharma et al (33) and Noureen et al (30) at 30 minutes, and Chamberlain et al (15), and Kapur et al (16) at 60 minutes.

**Figure 3:**
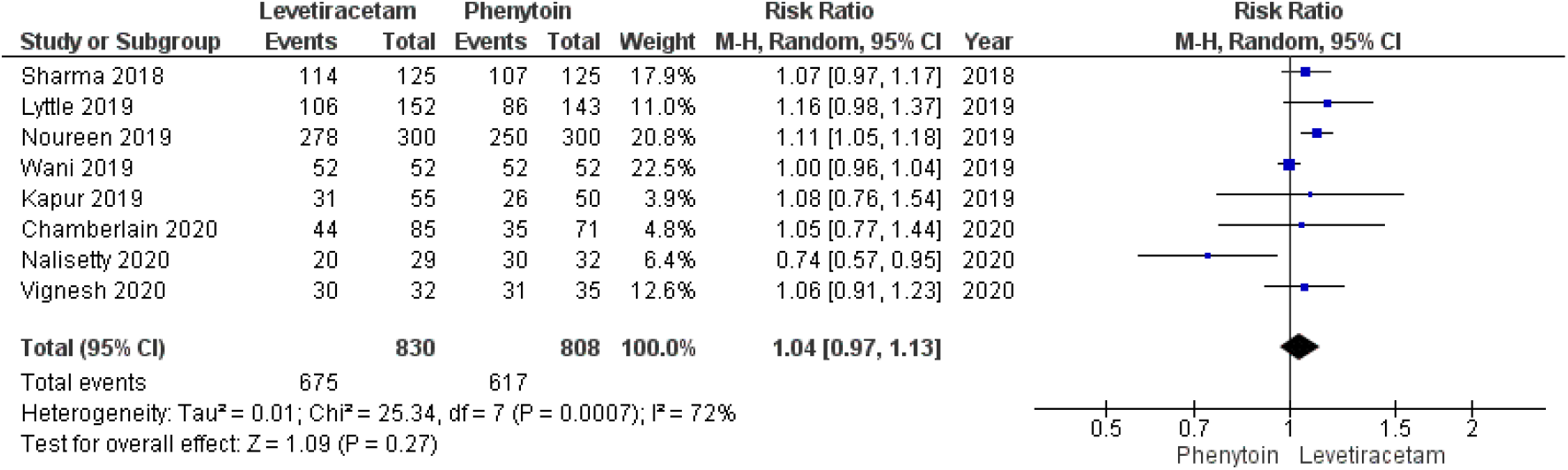
Seizure cessation within 5 min to 60 minutes of completion of study drug infusion.

#### Seizure recurrence between 1 hour to 24 hours of study drug infusion (Figure 4)

Seizure recurrence between 1-24 hours and/or use of third-line AEDs was reported by 8 RCTs (15, 16, 18, 20, 31-34). In the metanalysis, 8.9% (52/578) children in the LEV group and 15.1% (81/534) in the phenytoin group had seizure recurrence between 1 to 24 hours [RR 0.64 (95% CI 0.42-0.99), p=0.04] (figure 4) with no statistical heterogeneity (I^2^ of 28% and p=0.21).

**Figure 4:**
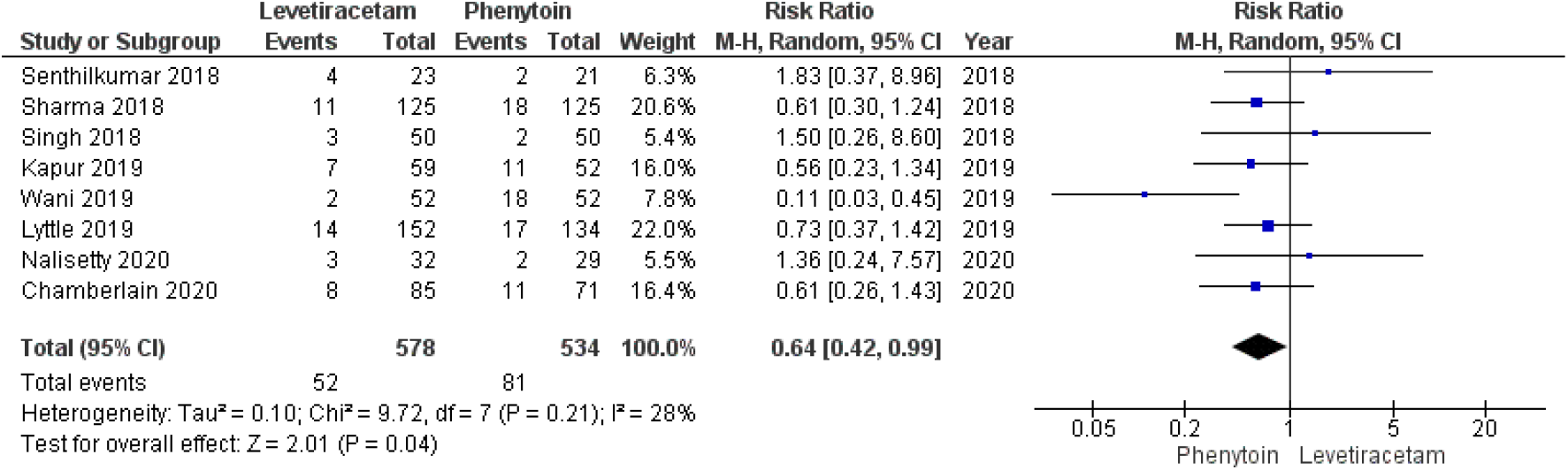
Seizure recurrence with in 1 hour to 24 hours of study drug infusion.

#### Time to achieve seizure cessation (Figure 5)

Time to achieve seizure cessation from following the study drug infusion was reported by 5 RCTs (19, 20, 31-33). The time [in minutes as mean (1)] to achieve seizure cessation was similar in both LEV group and PHT group [RR 0.49 (95% CI of 0.10-1.08), p=0.11] (Figure 5).

**Figure 5:**
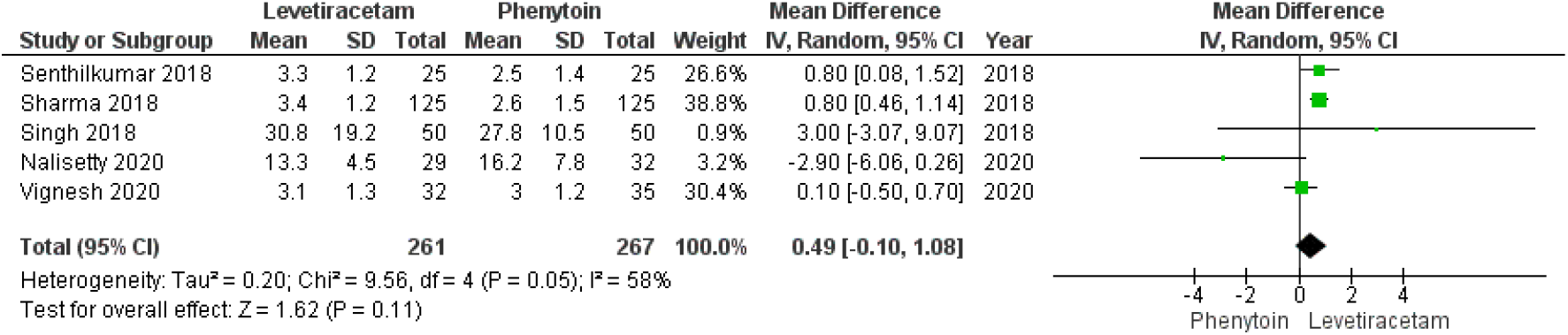
Time to achieve seizure cessation following completion of study drug infusion.

### Safety analysis (Figure 6 and 7)

Intubation/mechanical ventilation was required in 15% (103/674) children in the LEV arm and 23% (141/616) in the PHT arm (15, 17-20, 33). Although higher number of children in the PHT arm required intubation/mechanical ventilation, the difference was not statistically significant [RR 0.56 (95% CI 0.30-1.03), p=0.06] with marked heterogeneity among these RCTs (I^2^ 80%, p=0.0001) (Figure 6).

**Figure 6:**
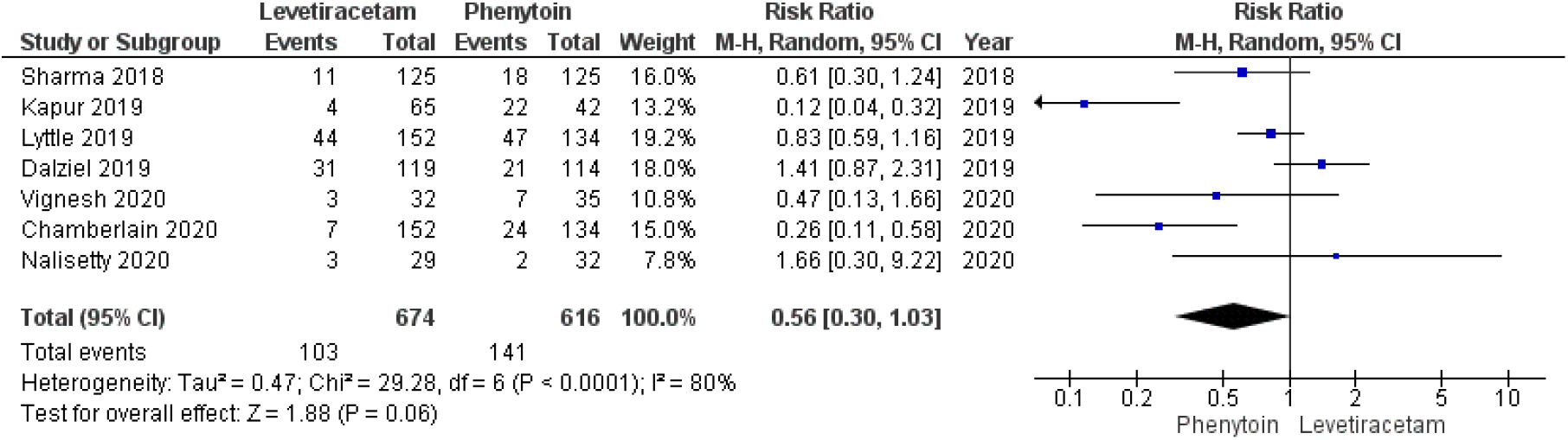
Requirement of intubation/or mechanical ventilation in the study groups after completion of study drug infusion.

**Figure 7:**
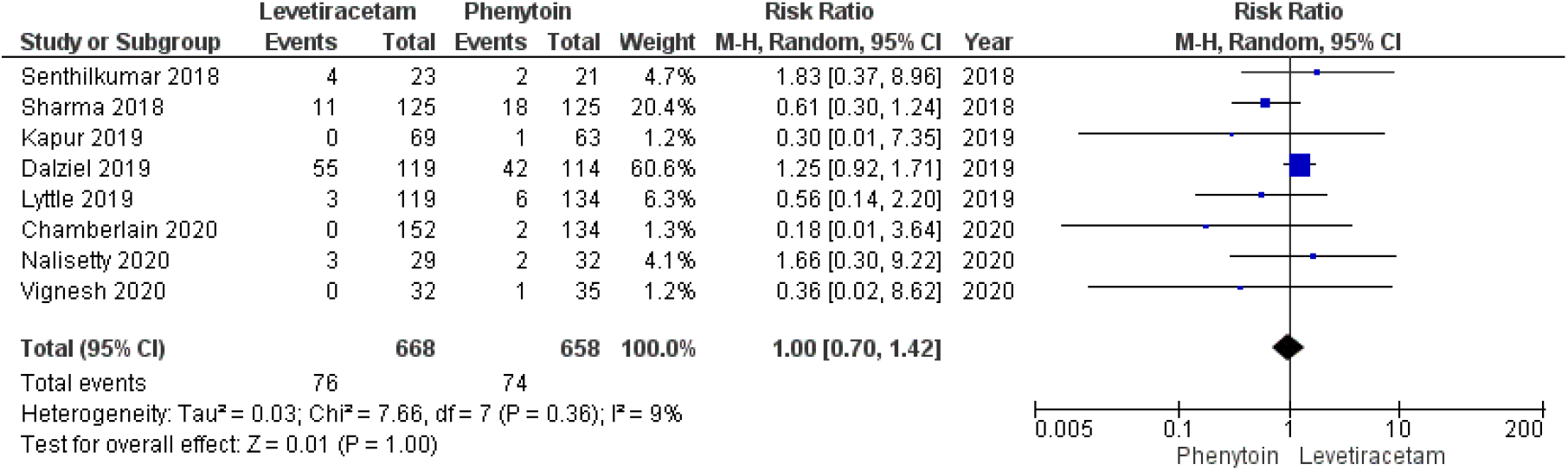
Cardiovascular instability (hypotension and cardiac arrhythmia) in children with convulsive SE following LEV and phenytoin infusion.

Cardiovascular instability was reported in 11.3% (76/668) children in the LEV arm, and 11.2% (74/658) in the phenytoin arm [RR 1, 95% CI 0.7-1.42, p=0.36] with homogenous RTCs (I^2^ 9%, and p=0.36) (15, 17-20, 32, 33) (Figure 7).

A total of 4 children died among 2019 total enrolled with mortality rate of 0.1%, two each died in LEV and PHT group. PICU admission and duration of hospital stay was not uniformly reported in the selected RCTs.

#### Risk of bias

The funnel plot for the RCTs reporting outcome of seizure cessation within 5 min to 60 minutes of completion of study drug infusion is shown in Figure 8. Three RCTs had low risk of bias (15, 16, 19) whereas 8 RCTs had high risk of bias (Figure 6a, 6b) (17, 18, 20, 30, 32-34). The most common bias observed were lack of blinding of primary outcome assessor, patients, parents and study team members. The blinding was also compromised by the study drug infusion methods, where there was difference in the study drug infusion time, adding to potential source of bias. The other bias was lack of allocation concealment in one study, small sample size, and reporting seizure cessation at various time points in one study (Figure 9 and 10).

**Figure 8:**
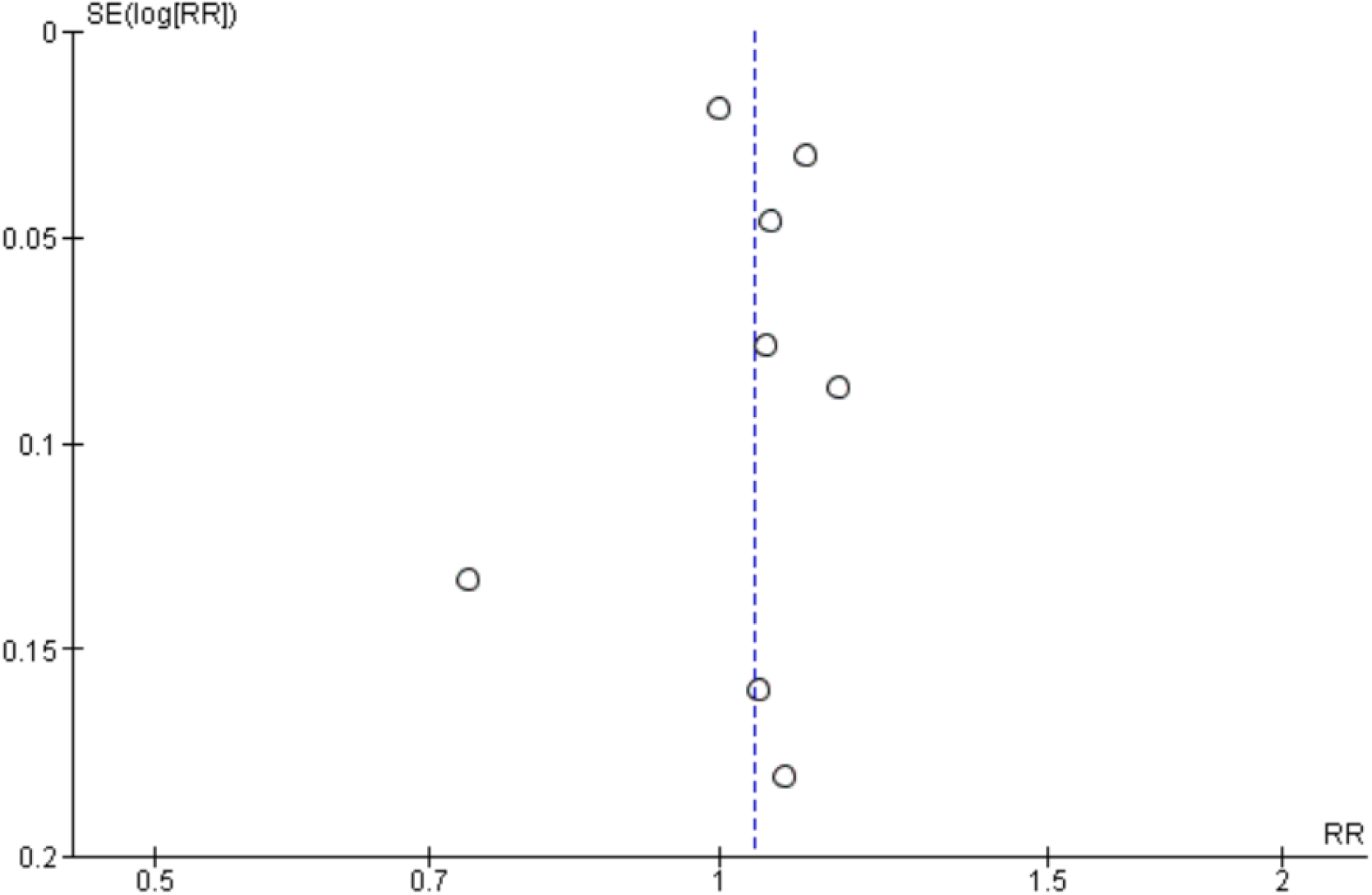
Funnel plot for the RCTs reporting outcome of seizure cessation within 5 min to 60 minutes of completion of study drug infusion.

**Figure 9:**
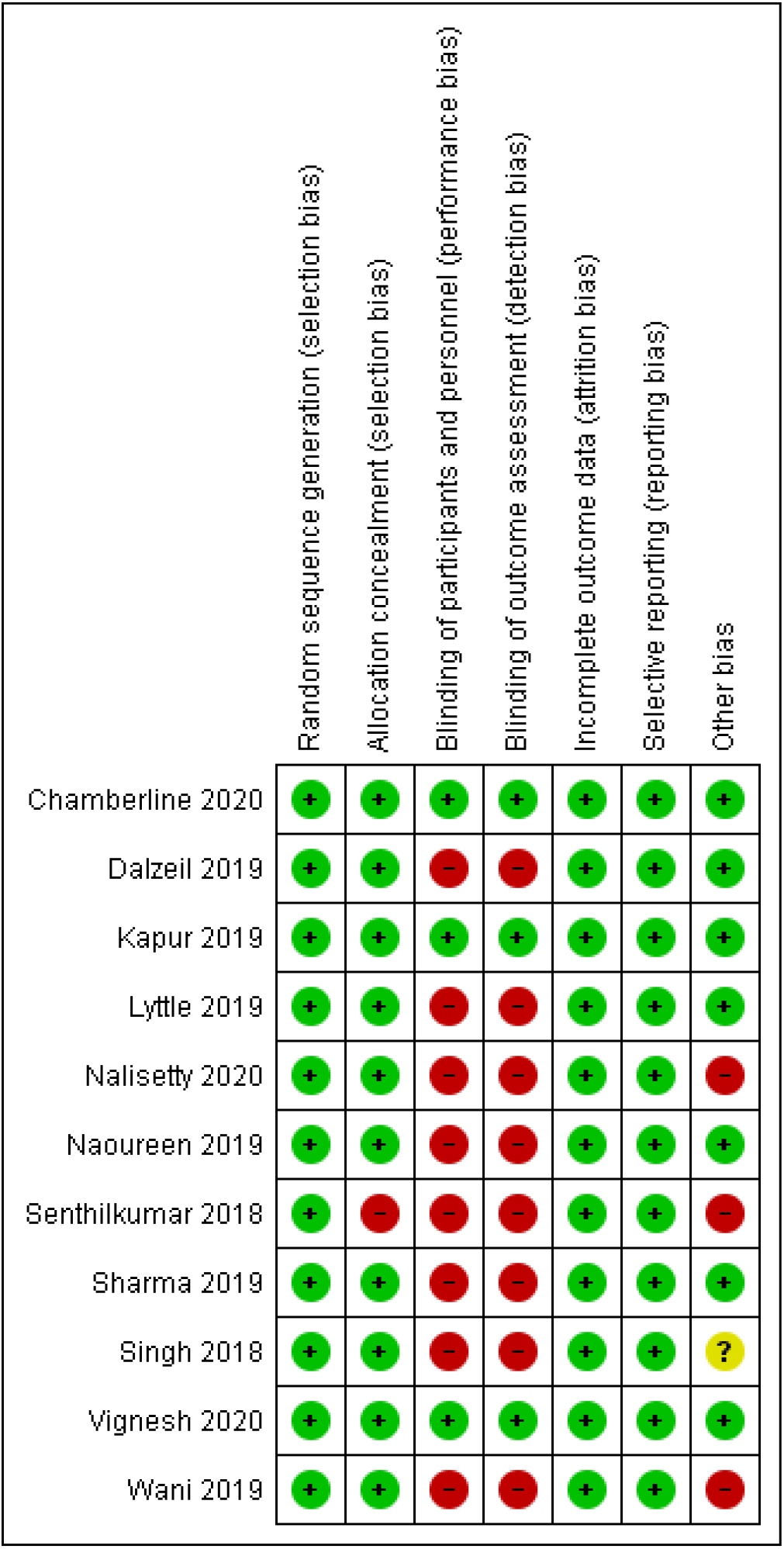
Risk of bias summary of individual RCTs assessed through Cochrane risk of bias tool.

**Figure 10:**
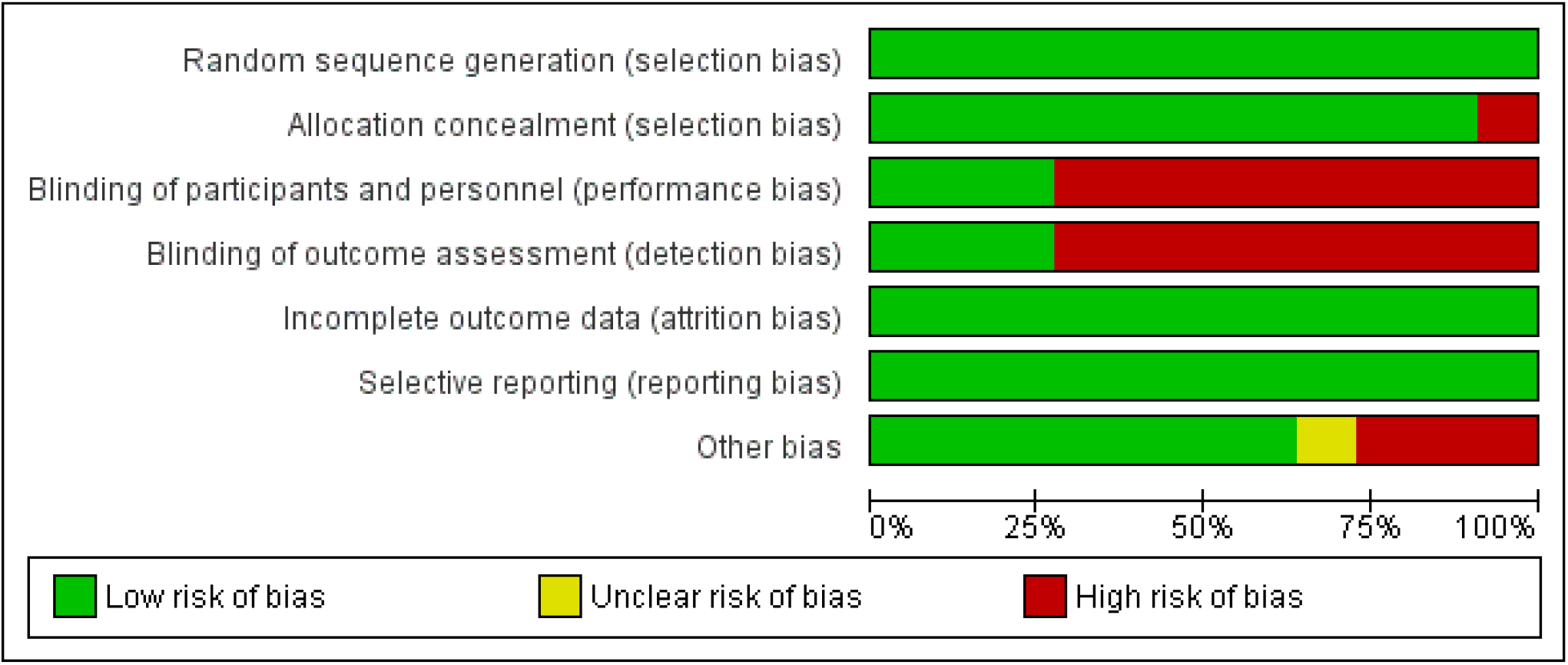
Risk of bias graph of all enrolled RCTs.

## Discussion

In this systemic review and meta-analysis of 11 RCTs involving 2177 children, we demonstrated that the LEV was not superior to PHT as second line agent for Pediatric convulsive SE. The efficacy of LEV and PHT varied from 50-94% and 49-80%, respectively (15, 17, 19). In the pooled analysis, efficacy of LEV and PHT to control seizures within 5 minutes of study drug infusion (61% vs. 65%), 5 minutes to 1 hour of study drug infusion (81% vs. 76%) were similar, respectively. The time to achieve seizure cessation was also similar in both the groups. However, the seizure recurrence rate within 24 hours of study drug infusion was significantly higher in PHT group as compared to LEV. The rate of cardiovascular instability was similar with both the groups whereas the rate of intubation/RSI or mechanical ventilation was higher with PHT group but was not statistically significant.

Both PHT and LEV were able to control seizures in 75-80% of case within 24 hours of study drug infusion. Maio et al (25) in a systematic review reported similar efficacy of LEV and PHT for seizure cessation and seizure recurrence within 24 hours for combined Pediatric and adult convulsive SE. Similarly, DeMott et al (24) reviewed 9 studies including adults with convulsive SE and reported that the efficacy of LEV and PHT was 74% vs 71%, respectively.

In this systematic review, 11% cases experienced cardiovascular instability and about 22% required intubation and mechanical ventilation following study drug infusion. The requirement of respiratory support was higher in PHT group, though not statistically significant. Lyttle et al (18) reported 30% children required intubation and mechanical ventilation in LEV group and 35% in PHT arm and two serious adverse events with hypotension and increased encephalopathy. Agitation was reported in 11% children who received LEV and 3% with phenytoin infusion (18). Kapur et al (16) reported acute respiratory depression in 12.8% in PHT group and 12% in LEV group. There was no significant difference between two drugs measured by episode of life-threatening hypotension and cardiac arrhythmia. Purple glove syndrome and acute anaphylaxis was not observed in these 11 selected RCTs.

In a systematic review of RCTs involving the adults with convulsive SE, the rates of respiratory depression were cardiac instability and mortality in PHT and LEV groups were 13% and 13%; 1.9% and 0.4%; 1.8% and 1.9%, respectively (24). Children experienced higher adverse events in comparison to adults with convulsive SE with both the drugs, however the rate were similar with both the drugs. One advantage of LEV observed was ease of infusion, as LEV was infused rapidly within 5-10 minutes (26). The dose of LEV varied considerably (20-60 mg/kg/dose), however in the absence of serious adverse events dosage upto 60 mg/kg dose can be used. Though numbers of hallucinations episodes reported were higher with 60 mg/kg LEV (16, 18).

It is important to promptly identify, treat, and investigate the SE in children, because SE lasting >30 minutes is associated with permanent neurological insult. Implementation of SE management protocols reduces the time to administer the ASM (35). Several SE management guidelines have been published to streamline management of pediatric SE (2, 11). For second line ASM between 20-40 minutes, there is no preferred single therapeutic agent among PHT, LEV and valproate (36). PHT is preferred second line ASM in management algorithms because of vast experience and familiarity with its use among emergency physicians (2, 11, 12). Similarly, for the third line ASM, there is no evidence-based single therapeutic agent, however midazolam, thiopentone, pentobarbitone or propofol are suggested to be used by 40-60 minutes of convulsive SE (2).

These management algorithms are modified at various levels based on the local practices and availability of PICU facility. In most of Pediatric SE guidelines, PHT is preferred second line agent, followed by midazolam infusion, if facility for PICU available, however if the facility for PICU is not available then valproate, phenobarbitone, and LEV are used in sequence, followed by the third line agent (6, 11, 37).

In a recent SE management in emergency pathway evaluation in 10 pediatric research centre across USA, fosphenytoin was used as first choice second line agent and PHT, LEV, valproate, and phenobarbitone were used as other second line agents, and several centres preferred use of additional bolus dose of AEDs before embarking upon the continuous midazolam or pentobarbitone infusion (36).

The strengths of this meta-analysis include the rigorous search strategy to include all RCTs that reported children with SE where PHT and LEV were tested as second line ASM. The efficacy and safety of both the drugs were evaluated. As the quality of meta-analysis depends on the quality of studies included. In this systematic review and meta-analysis, the quality of 8 studies was compromised due to lack of allocation concealment, blinding, and small sample size. The primary outcome observer, patient, and study personnel were not blinded to the study drug. Second limitation of this systematic review was the different primary and secondary outcome parameters used by the authors for the efficacy analysis. There is marked heterogeneity among the included studies.

The dose of LEV varied considerably among the studies; ranging from 20-60 mg/kg and the duration of infusion also varied. In the conSEPT trial LEV was infused within 5 minutes and phenytoin was infused at 20 minutes and outcome was assessed at 10 min and 25 minutes in respective arms (17). However, it is unclear with rapid infusion the LEV can cross the blood brain barrier too, as seizure cessation was achieved in 61% at 5 minutes and in 81% children by 1 hour. The assessment of seizure cessation was clinically in all studies and EEG was not assessed. Few patients might continue to have non-convulsive SE manifesting as lack of improvement in consciousness. Therefore, whenever available, EEG/vEEG monitoring would give more robust data for seizure control. However, the two RCTs (15, 16) with low risk of bias and using 60 mg/kg LEV dose, also reported similar efficacy and safety of LEV and PHT for Pediatric SE.

## Conclusion

Both LEV and PHT were equally effective in controlling Pediatric SE as second line ASM. However, PHT group has higher rate of seizure recurrences by 24 hours. The requirement of mechanical ventilation and cardiovascular instability were similar with both the drugs.

## Data Availability

Data will be available

## Figures and tables legends

**Table 1:** Study characteristics.

