## Supplementary material for "Efficacy and safety of levetiracetam versus phenytoin as second line antiepileptic agent in Pediatric convulsive status epilepticus: A systematic review and meta-analysis of randomized controlled trials": Table 1

**Table 1: Study details**

| S. no | Author, year | Country | No. of centres | No. of cases <18 years | Study duration | Age group enrolled | Levetiracetam | | | Phenytoin | | |
| --- | --- | --- | --- | --- | --- | --- | --- | --- | --- | --- | --- | --- |
|  |  |  |  |  |  |  | **Dose**  **(mg/kg)** | **Time to infusion** | **% and number of subjects achieving seizure cessation in 60 min** | **Dose**  **(mg/kg)** | **Time to infusion** | **% and number of subjects achieving seizure cessation in 60 min** |
| 1 | Chamberlain et al^15^, 2020 | USA | 58 | 225 | Nov 2015 to Dec 2018 | >2 years | 60 | 10min | 51% (44/85) | 20 | 10min | 49% (35/71) |
| 2 | Vignesh et al^19^, 2020 | India | 1 | 110 | June 2016 to Dec2018 | 3 months-12 years | 20 | 20min | 94% (30/32) | 20 | 20min | 88% (31/35) |
| 3 | Nalisetty et al^20^, 2020 | India | 1 | 61 | June 2014 to Dec 2015 | 2 months-18 years | 40 | 10min | 69% (20/29) | 20 | 10 min | 94% (30/32) |
| 4 | Dalziel et al^17^, 2019 | Newzealand Australia | 13 | 233 | March 2015 to Nov 2017 | 3 months-16 years | 40 | 5 min | 50% (60/119) | 20 | 20 min | 60% (68/114) |
| 5 | Lyttle et al^18^, 2019 | UK | 30 | 286 | July 2015 to April 2018 | 6 month-18 years | 40 | 5 min | 70% (106/152) | 20 | 20 min | 60% (86/143) |
| 6 | Kapur et al^16^, 2019 | USA | 57 | 158 | Nov 2015 to Oct 2017 | >2 years | 60 | 10min | 56% (31/55) | 20 | 10min | 52% (26/50) |
| 7 | Noureen et al^30^, 2019 | Pakistan | 1 | 600 | Jan 2014 to June 2018 | 1-14 years | 40 | 15min | 93% (278/300) | 20 | 30 min | 83% (250/300) |
| 8 | Wani et al^34^, 2019 | India | 1 | 104 | June 2017- | 1 month -12 years | 40 | 10 min | 100% (52/52) | 20 | 20 min | 100% (52/52) |
| 9 | Singh et al^31^ 2019 | India | 1 | 100 | Nov 2012 to April 2014 | 3-12 years | 30 | 6 min | 94% (47/50) | 20 | 20 min | 96% (48/50) |
| 10 | Senthil Kumar et al^32^, 2019 | India | 1 | 50 | Jan 2017-June 2017 | 3 months-12 years | 30 | 7 min | 92% (23/25) | 20 | 7 min | 84% (21/25) |
| 11 | Sharma et al^33^, 2019 | India | 1 | 250 | March 2017 to September 2018 | 6 months-18 years | 20 | - | 91% (114/125) | 20 | - | 86% (107/125) |
